# Delay Discounting and Family History of Psychopathology in Children Ages 9-11: Results from the ABCD Study

**DOI:** 10.1101/2023.07.20.23292967

**Authors:** Matthew E. Sloan, Marcos Sanches, Jody Tanabe, Joshua L. Gowin

**Affiliations:** Addictions Division, Centre for Addiction and Mental Health, Toronto, Ontario, Canada; Department of Pharmacology & Toxicology, University of Toronto, Toronto, Ontario, Canada Canada; Division of Neurosciences and Clinical Translation, Department of Psychiatry, University of Toronto, Toronto, Ontario, Canada; Campbell Family Mental Health Research Institute, Centre for Addiction and Mental Health, Toronto, Ontario, Canada; Department of Psychological Clinical Science, University of Toronto Scarborough, Toronto, Ontario, Canada; Institute of Medical Science, University of Toronto, Toronto, Ontario, Canada; Biostatistics Core, Centre for Addiction and Mental Health, Toronto, Ontario; University of Colorado Anschutz Medical Campus, Department of Radiology, Aurora,□CO□80045

**Keywords:** Impulsivity, Development, Cognition, Risk, Depression, Mania, Alcohol, Substance Use

## Abstract

Delay discounting is a tendency to devalue delayed rewards compared to immediate rewards. Evidence suggests that steeper delay discounting is associated with psychiatric disorders across diagnostic categories, but it is unclear whether steeper delay discounting is a risk factor for these disorders. We examined whether children at higher risk for psychiatric disorders, based on family history, would demonstrate steeper delay discounting behavior. We examined the relationship between delay discounting behavior and family history of psychopathology using data from the Adolescent Brain Cognitive Development (ABCD) study, a nationally representative sample of 11,878 children. Participants completed the delay discounting task between the ages of 9 and 11. We computed Spearman’s correlations between family pattern density of psychiatric disorders and delay discounting behavior. We conducted mixed effects models to examine associations between family pattern density of psychiatric disorders and delay discounting while accounting for sociodemographic factors. Correlations between family history of psychopathology and delay discounting behavior were small, ranging from ρ = –0.02 to 0.04. In mixed effects models, family history of psychopathology was not associated with steeper delay discounting behavior. Sociodemographic factors played a larger role in predicting delay discounting behavior than family history of psychopathology. Race, ethnicity, sex, parental education, and marital status were all significantly associated with delay discounting behavior. These results do not support the hypothesis that children with greater risk for psychopathology display steeper delay discounting behavior. Sociodemographic factors play a larger role in determining delay discounting behavior in this age group.

## 1.1 Introduction

Delay discounting refers to the tendency to devalue delayed rewards compared to more immediately available rewards (Bickel and Marsch 2001). The longer the delay, the less an individual tends to value the reward (Kirby 1997; Madden et al. 2003). Delay discounting is conserved across species, including pigeons, rats, and non-human primates (Vanderveldt et al. 2016). Among humans, delay discounting behavior can differ substantially between individuals. Differences in delay discounting may reflect choice impulsivity, with more impulsive individuals prone to seek immediate rewards at a higher discount than less impulsive individuals (Ainslie 1975; Levitt et al. 2020). There is evidence that choice impulsivity does not correlate with other measures of impulsivity but represents a unique construct (Hamilton et al. 2015). Delay discounting tasks have been used to study differences in choice impulsivity between individuals with and without psychiatric disorders and delay discounting has been proposed as a treatment target for some disorders (Koffarnus et al. 2013).

There is evidence that steeper delay discounting may be a transdiagnostic process in psychiatric disorders. In support of this, a meta-analysis found that steeper discounting was observed across a range of diagnoses including major depressive disorder, bipolar disorder, borderline personality disorder, obsessive compulsive disorder, and bulimia nervosa (although individuals with anorexia nervosa exhibited shallower delay discounting compared to controls) (Amlung et al. 2019). Other studies have found delay discounting to be steeper in individuals with substance use disorders and addictive behaviors (Amlung et al. 2017; Gowin et al. 2018). However, the utility of delay discounting has been questioned, as delay discounting behavior is only modestly correlated with measures of psychological dysfunction and does not have adequate sensitivity or specificity for any particular disorder (Bailey et al. 2021).

Several important elements of the relationship between delay discounting and psychopathology remain unclear. First, although there are differences between cases with psychiatric disorders and healthy controls, it is unknown whether steeper delay discounting is a risk factor for psychiatric disorders or represents a manifestation of the disorder itself. Also, since most delay discounting tasks examine monetary choices, it is possible that socioeconomic differences between cases and controls could explain a substantial amount of the variance in delay discounting behavior between groups. For example, in a previous analysis of differences in delay discounting between individuals with and without alcohol use disorder, we found that education and household income explained a greater proportion of the variance in delay discounting behavior than diagnostic group (Gowin et al. 2019). Globally, better financial environments have been associated with shallower delay discounting behavior whereas inflation and inequality have been associated with steeper rates of discounting (Ruggeri et al. 2022). Data from large prospective studies with participants from a range of socioeconomic backgrounds offer an opportunity to improve our understanding of the link between delay discounting behavior and future psychopathology.

In order to better understand whether delay discounting represents a risk factor for psychiatric illness, we analyzed data from the Adolescent Brain Cognitive Development (ABCD) study. The ABCD study is one of the largest developmental studies ever undertaken, with prospective data collected from over 11,000 children (Casey et al. 2018). Because ABCD measured delay discounting prior to the age of onset of many psychiatric disorders, this dataset is uniquely positioned to look at the association between delay discounting behaviors and risk of psychiatric illness. Since most psychiatric disorders are highly heritable, we used family history of psychiatric illness as an index of risk in our analyses. We hypothesized that steeper delay discounting would be associated with risk of psychiatric illness with a moderate effect size. As ABCD collected extensive socioeconomic and demographic data from participants, we also examined the association between these variables and delay discounting behavior with the hypothesis that socioeconomic status would account for some but not all of the association between family history of psychiatric illness and delay discounting behavior.

## 2.1 Materials and Methods

### 2.1.1 Sample

The sample for the ABCD study consists of 11,878 participants; one goal of the study was to have sufficient power to detect small to medium effects (Garavan et al. 2018). The study recruited participants from 21 sites in the U.S., including 4 sites from the northeast, 4 from the Midwest, 6 from the south, and 7 from the west (Garavan et al. 2018). Recruitment for the ABCD study launched in September 2016 and initially recruited children aged 9-10. For this analysis, we accessed data from release 4.0. We used data from the baseline assessment, when data about family history of psychopathology was collected, as well as data from the one year follow up assessment, when the delay discounting task was first administered. A previous paper has examined family history of harmful alcohol use (Kohler et al. 2022), but no previous reports have examined family history of other mental health issues.

## 2.2 Delay Discounting Task

In the ABCD study’s delay discounting task, participants make a series of choices between two monetary options with different delays (e.g., $80 now or $100 three months from now) (Koffarnus and Bickel 2014). Seven delay periods were tested: 6 hours, 1 day, 1 week, 1 month, 3 months, 1 year, and 5 years. Each delay period included six trials and the choices offered were contingent on prior choices. For example, if the participant chose the immediate amount on trial 1, the immediate amount offered was decreased on trial 2. By contrast, if they chose the delayed amount, the immediate amount was increased. The six trials are used to establish an indifference point, which is the amount of money received immediately that is equivalent to $100 after a specified delay for that person (e.g., $80 now = $100 in 1 month). Using the indifference points at the seven delay periods, there are several methods to generate a summary metric of delay discounting. The most common metrics are the area under the indifference point versus delay curve and the delay discounting constant, k, which is a parameter in a hyperbolic function to describe how steeply an individual discounts commodities across time. Area under the curve and k are anti-correlated, where lower values of area under the curve, but higher values of k, represent steeper delay discounting. Our primary metric here is the unweighted area under the curve, which places equal weight on the 7 indifference points. We also report a weighted area under the curve, where the indifference point is multiplied by the length of the delay, as well as the k parameter and its natural log, since the distribution of k is skewed.

In order to retain only higher quality data, an algorithm has been developed to identify cases where indifference points do not monotonically decrease with increasing delays (Johnson and Bickel 2008). According to this algorithm, data are classified as nonsystematic if (1) “any indifference point (starting with the second delay) was greater than the preceding indifference point by a magnitude greater than 20% of the larger later reward” and (2) “if the last… indifference point was not less than the first… indifference point by at least a magnitude equal to 10% 0f the larger later reward” (Johnson and Bickel 2008). Our secondary analyses only retained individuals who met these data quality criteria.

## 2.3 Family History of Psychopathology

Family history of psychopathology was measured at the baseline ABCD assessment using the Family History Assessment Module Screener (FHAM-S) from the NCANDA study (Brown et al. 2015; Rice et al. 1995). The 11-item FHAM-S asks caregivers questions screening for a family history of alcohol and drug problems, depression, mania, schizophrenia, and suicidality in first and second-degree relatives (i.e., parents, siblings, grandparents, uncles, and aunts). Single questions are used for each issue. For example, to assess for alcohol problems, the FHAM-S asks “has drinking ever caused any of your relatives to have problems with health, family, job, or police?” (https://niaaagenetics.org/coga_instruments/phaseI/fham/Fhamhist4-scr.pdf). For each question, the caregiver lists the name and relationship to the proband of each relative who they endorse as having the specified problem. In our primary analyses, we did not use the questions about whether relatives got into trouble with the police from time to time, ever had problems with their nerves or a nervous breakdown, ever talked to a doctor or counselor about emotional or mental problems, or ever had been hospitalized because of emotional or mental problems as these items were not felt to be specific to any one disorder.

To calculate a score for family history of psychopathology, we used family pattern density of each disorder as our primary metric (Ranganathan et al. 2014). Family pattern density uses data from all relatives and takes into account how genetically distant each affected relative is from the proband when calculating the summary score, which ranges from 0 to 3 (Ranganathan et al. 2014). In this method, each affected parent would count for a half point and each grandparent counts for a quarter point. Siblings (= 0.5 base value), uncles (= 0.25 base value), and aunts (= 0.25 base value) are rated in a way that is proportional to their number and genetic relatedness (e.g. if there are two paternal uncles and one has the problem in question, they would count for 0.25/2 = 1/8 points). Other methods for calculating family history of psychopathology were used in our sensitivity analyses, such as family history density (Stoltenberg et al. 1998), which only uses parents (rated at half a point for each family history positive parent) and grandparents (rated at a quarter point for each family history positive grandparent), with scores ranging from 0 to 2, and family patterns analysis (Turner et al. 1993), which assigns positive parents and grandparents a score of 1 whereas siblings, aunts, and uncles are given a score of one divided by the number of siblings, aunts, or uncles (so 2 out of four siblings with a problem would generate a score of 2/4 or 0.5).

## 2.4 Sociodemographic Measures

Age, sex, race, ethnicity, household income, parental working status, parental marriage status, and parental education level were used as covariates in our adjusted analyses. Ethnicity refers to whether the individual identified as Hispanic or not. We used four racial categories: White, Black, Asian, or Other/Mixed. These variables were also used in an additional analysis looking at the effects of these variables on delay discounting without controlling for family history of psychopathology.

## 2.5 Statistical Analysis

Our primary analyses included all individuals who completed the delay discounting task. We corroborated these results in the subsample that met data quality criteria for the delay discounting task. We assessed Spearman’s correlations between family pattern density of each problem (alcohol, drug, depression, mania, schizophrenia, and suicide) and delay discounting outcomes [AUC, weighted AUC, hyperbolic AUC, and ln(k)]. We chose to use Spearman’s correlations because family pattern density data was highly skewed. Next, we used mixed effects models to determine whether there were associations between family pattern density of each problem and delay discounting AUC when accounting for the sociodemographic variables listed above. Levels in each model included the individual (level 1), family (level 2) as some participants were siblings within the same family, and study site (level 3). Family and study site were specified as random intercepts in the model. Since we examined six family history variables (alcohol problems, drug problems, depression, mania, schizophrenia, suicidal behavior), we applied a Bonferroni correction and used alpha ≤ 0.008 as our threshold for statistical significance. All analyses were conducted using R.

We conducted numerous sensitivity analyses. We repeated all analyses using family history density and family patterns analysis scores. We also conducted primary analyses in the full sample and repeated them in the sample that met data quality criteria for delay discounting task (Johnson and Bickel 2008). Results from secondary analyses can be found in the supplementary materials.

## 3.1 Results

### 3.1.1 Sample Characteristics

There were a total of 11,876 participants in the sample. Of these, 11,065 participants had complete delay discounting data at the 1-year follow up. Less than half of the sample (N = 4,364 or 39.4% of the sample) met data quality criteria for the delay discounting task (Johnson and Bickel 2008) (Table 1). There were significant demographic differences between the full sample and the subsample that met data quality criteria, where those that met the criteria were more likely to be white, non-Hispanic, have higher household income, greater parental education, and live in houses where their parents were married (Table 1). There were also significant correlations among family history variables (Supplemental Figure 1), with the greatest associations between history of alcohol and drug problems (ρ = 0.43), alcohol problems and depression (ρ = 0.29), depression and suicidal behavior (ρ = 0.33), and depression and mania (ρ = 0.29).

**Table 1.** Demographic data for the sample that met and did not meet the data quality criteria for the delay discounting task. There were significant differences between groups (p < 0.001) for all sociodemographic variables included. Missing data in the overall sample (N, %): marital status of parents (73, 0.7%), household income (897, 8.1%), education level (11, 0.1%), ethnicity (62, 0.6%), and race (151, 1.4%).

|  | Met Data Quality Criteria | Did Not Meet Data Quality Criteria |
| --- | --- | --- |
| **Participants [N (%)]** | 4364 (39.4%) | 6,701 (60.5%) |
| **Area Under the Indifference Point Curve (AUC)** | 69.0 (20.4) | 64.7 (24.7) |
| **Ln(k)** | -1.8 (2.3) | -2.1 (4.4) |
| **Fam. Hist. Density of Alcohol** | 0.19 (0.32) | 0.20 (0.34) |
| **Fam. Hist Density of Drug** | 0.11 (0.25) | 0.13 (0.30) |
| **Fam. Hist Density of Depression** | 0.34 (0.48) | 0.34 (0.48) |
| **Fam. Hist Density of Schizophrenia** | 0.02 (0.09) | 0.03 (0.11) |
| **Fam. Hist. Density of Mania** | 0.04 (0.15) | 0.05 (0.16) |
| **Fam. Hist. Density of Suicidal Behavior** | 0.05 (0.15) | 0.05 (0.16) |
| **Age [Mean (SD)]** | 10 (0.6) | 9.8 (0.6) |
| **Sex [Female N (%)]** | 1981(45.4%) | 3297 (49.2%) |
| **Parents Married [N (%)]** | 3265 (74.8%) | 4311 (64.3%) |
| **Household Income [N (%)]** | <\$50,000: 839 (19.2%)<br>>50K-100K: 1167 (26.7%)<br>>\$100,000: 2053 (47.0%) | <\$50,000: 2045 (30.5%)<br>>50K-100K: 1738 (25.9%)<br>>\$100,000: 2326 (34.7%) |
| **Education [N (%)]** | < HS/GED: 113 (2.6%)<br>HS/GED: 245 (5.6%)<br>Some College: 921 (21.1%)<br>Bachelor's: 1247 (28.6%)<br>Graduate: 1836 (42.1%) | < HS/GED: 407 (6.1%)<br>HS/GED: 745 (11.1%)<br>Some College: 1882 (28.1%)<br>Bachelor's: 1606 (24.0%)<br>Graduate: 2052 (30.6%) |
| **Hispanic (N [%])** | 633 (14.5%) | 1201 (17.9%) |
| **Race** | White: 3138 (71.9%)<br>Black: 386 (8.8%)<br>Asian: 115 (2.6%)<br>Other/Mixed: 686 (15.7%) | White: 4012 (59.9%)<br>Black: 1247 (18.6%)<br>Asian: 140 (2.1%)<br>Other/Mixed: 1190 (17.8%) |

## 3.2 Correlations Between Delay Discounting Behavior and Family History of Psychiatric Disorders

Correlations between family pattern density and delay discounting behavior (as measured by AUC) were small, ranging from –0.02 to 0.04 (Figure 1), indicating that family pattern density of psychiatric disorders accounted for less than 0.2% of the variance in delay discounting behavior. Only family history of depression (ρ = 0.04, p < 0.001) was significantly associated with unweighted AUC after applying Bonferroni correction. There were no statistically significant associations between family pattern density of alcohol problems, drug problems, mania, schizophrenia, or suicidal behavior and delay discounting behavior. For depression, the direction of the correlation was opposite to the hypothesized direction, with individuals who had a greater family pattern density of these disorders showing shallower delay discounting behavior. Findings were similar in the subsample meeting data quality criteria for the delay discounting task (Figure S2) or when using family history density or family patterns analysis instead of family pattern density (Figure S3 and S4).

**Figure 1.**
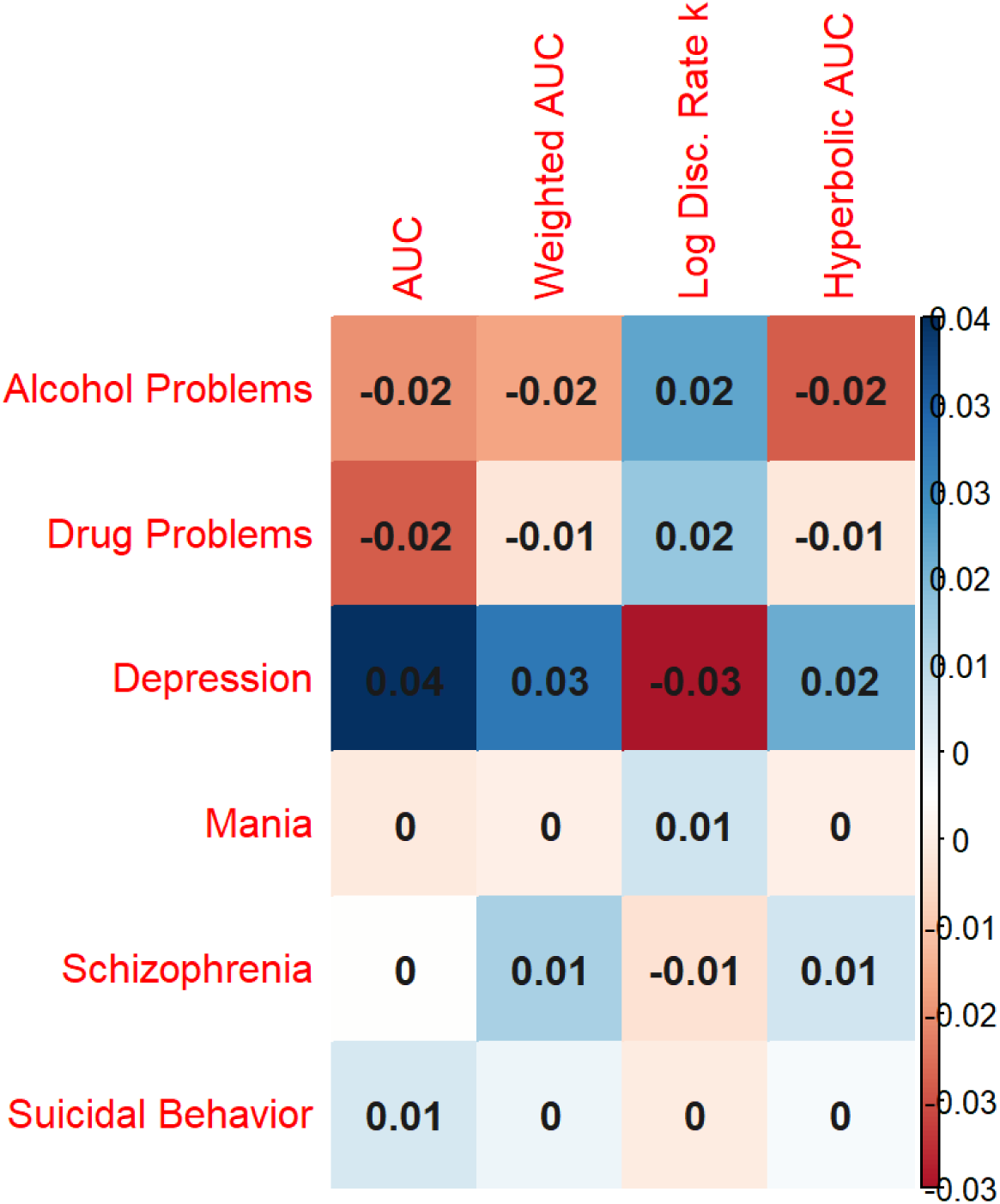
Spearman’s correlations between different delay discounting metrics and family history of alcohol problems, drug problems, depression, mania, schizophrenia, and suicide attempt or completion (measured using family pattern density scores). Correlations between family history of each disorder and the area under the indifference point versus delay curve were small in magnitude, ranging from – 0.02 to 0.04.

## 3.3 Associations Between Delay Discounting Behavior and Family History of Psychiatric Disorders in Mixed Effects Models

There were no associations between any family pattern density variable and delay discounting behavior in mixed effects models with unweighted AUC as the dependent variable after adjusting for co-variates and applying Bonferroni correction (Figure 2). Coefficient estimates for each disorder are as follows: alcohol problems (coefficient = –1.55, 95% CI = –2.88 to –0.22, p = 0.022), drug use problems (coefficient = –1.43, 95% CI = –3.04 to 0.19, p = 0.084), depression (coefficient = 0.13, 95% CI = –0.80 to 1.06, p = 0.787), mania (coefficient = –2.91, 95% CI = –5.69 to –0.14, p = 0.040), schizophrenia (coefficient = 0.62, 95% CI = –3.74 to 4.98, p = 0.780), and suicide attempt/completion (coefficient = –0.04, 95% CI – 2.82 to 2.73, p = 0.975) (full results of each model can be found in Figures S6 – S11 and Supplementary Tables S1 – S6). Results were similar in the subsample meeting data quality criteria for the delay discounting task (Figure S12; for full models in this sample see Figures S13 – S18 and Tables S7 – S12) or when using family history density or family patterns analysis instead of family pattern density (Figure S19 and S20).

**Figure 2.**
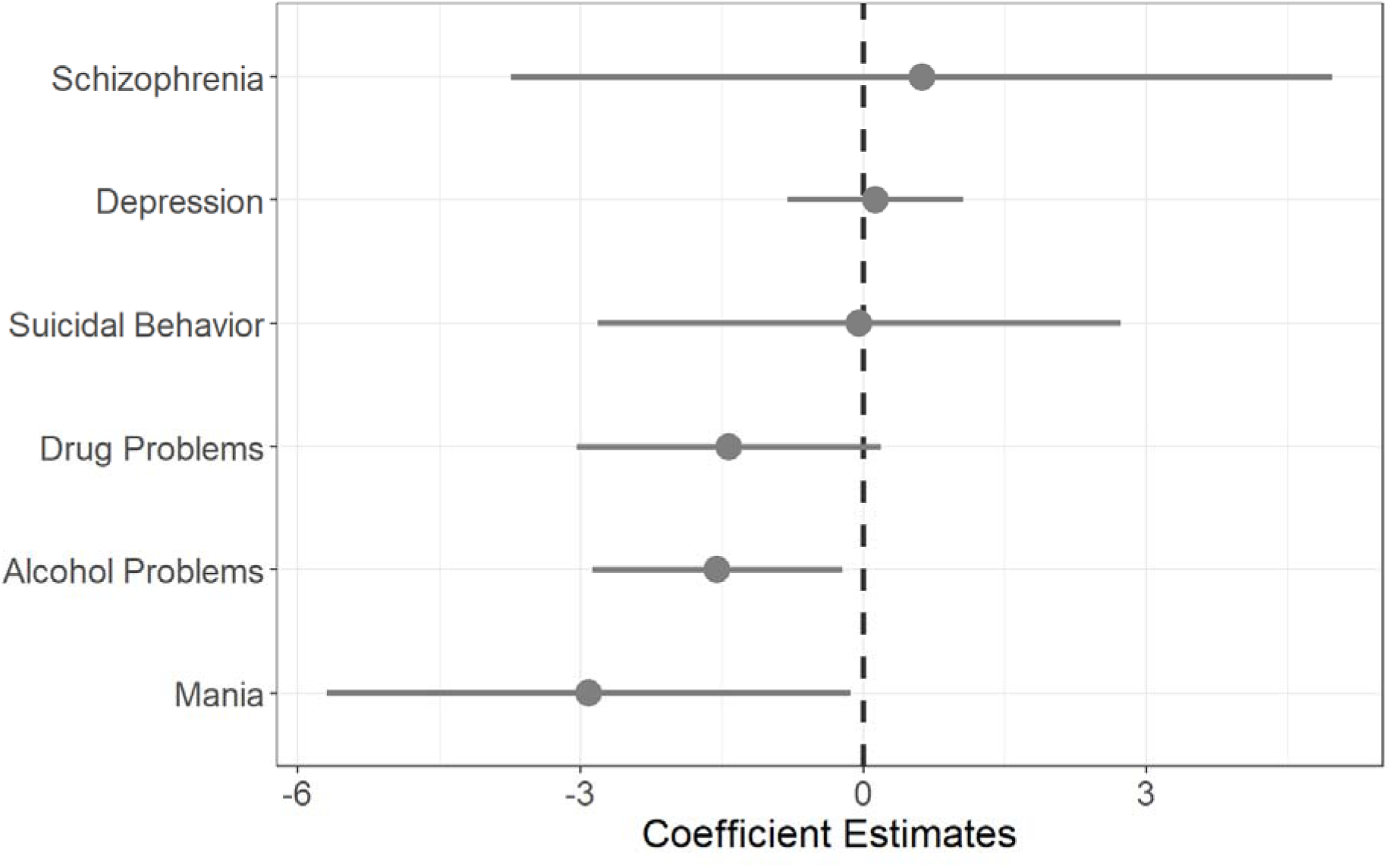
Mixed effects models (Tables S1-S6) did not find any significant associations between family history of psychiatric disorders and delay discounting behavior when adjusted for socioeconomic and demographic variables and applying Bonferroni correction. Error bars represent 95% confidence intervals for unstandardized coefficient estimates. Area under the indifference point versus delay curve is the dependent variable, so lower coefficient values indicate greater delay discounting.

## 3.4 Associations Between Delay Discounting Behavior and Sociodemographic Variables in Mixed Effects Models

Race, ethnicity (Hispanic or non-Hispanic), sex, parental education, and parental marital status were associated with delay discounting behavior in the above mixed effect models (Supplementary Tables S1-S6, Figures S6 – S11). The largest effect sizes were observed for parental education, with children whose parents had completed a bachelor’s degree (coefficient = 5.14, 95% CI 2.6 to 7.69, p < 0.001) or postgraduate education (coefficient = 6.29, 95% CI 3.71 to 8.86, p < 0.001) displaying shallower delay discounting behavior compared to those who had not completed high school. Children identified by their parent as Black had steeper delay discounting than children identified as White (coefficient = – 7.02, 95% CI –8.48 to –5.55, p < 0.001) and children identified as Hispanic had steeper delay discounting than children identified as non-Hispanic (coefficient = –3.88, 95% CI –5.28 to –2.49, p < 0.001). Males had steeper delay discounting than females (coefficient = –2.66, 95% CI –3.52 to –1.80, p < 0.001). Children whose parents were married had shallower discounting than children whose parents were separated or divorced (coefficient = 1.30, 95% CI 0.14 to 2.45, p = 0.028). Children from a household earning between $50,000 and $100,000 per year had shallower delay discounting relative to children from households earning less than $50,000 per year (coefficient = 1.70, 95% CI 0.30 to 3.10, p = 0.018). Age was not associated with delay discounting behavior. Results from a mixed effects model containing only sociodemographic variables can be found in Figure 3.

**Figure 3.**
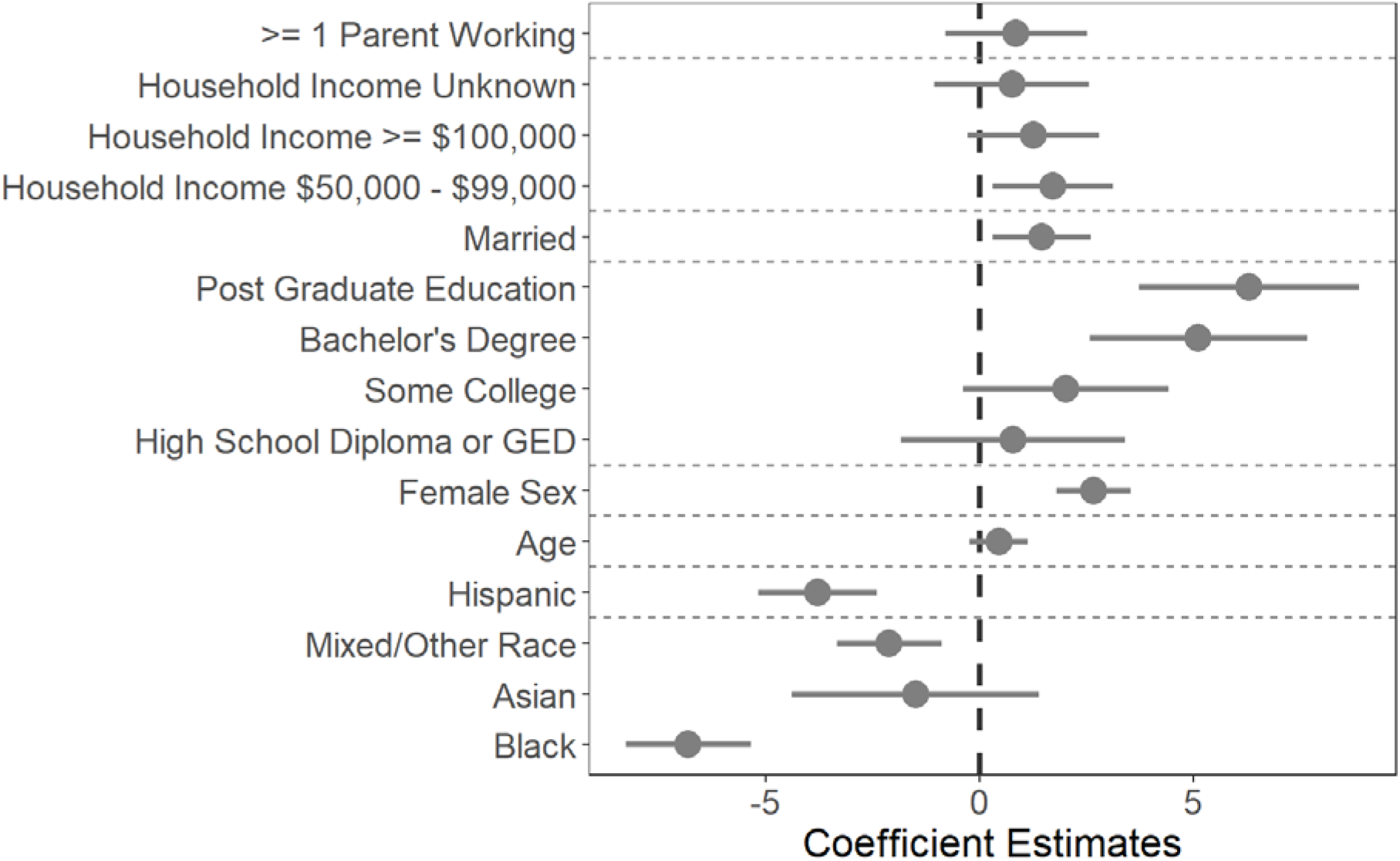
Reference groups were (variable of interest in brackets): household income < $50,000 (household income), unmarried (marital status), did not complete high school (education), White (race), non-Hispanic (ethnicity), male (sex). Age (years) is a continuous variable. Children whose parents were married and whose parents had a graduate or bachelor’s degree had shallower delay discounting than those whose parents were unmarried or had not completed high school. Females had shallower delay discounting behavior than males. Individuals who identified as Black or Hispanic had steeper delay discounting than those who identified as White or non-Hispanic. Error bars represent 95% confidence intervals. In this figure, the area under the indifference point versus delay curve is the dependent variable, so lower coefficient values indicate greater delay discounting.

## 4.1 Discussion

The results of our study suggest that children at higher risk of psychopathology do not display steeper delay discounting behavior at ages 9-11. For all disorders, correlations between family pattern density and delay discounting behavior were close to 0, indicating that family pattern density of each disorder accounted for only a small fraction of the variance in delay discounting behavior. In mixed effects models accounting for sociodemographic variables, there were no statistically significant associations between family pattern density of any psychiatric disorder and delay discounting behavior. By contrast, we found associations between delay discounting behavior and sociodemographic factors including parental education, parental marital status, sex, race, and ethnicity. Our findings suggest that at this age, sociodemographic factors have a greater impact on delay discounting behavior than family history of psychopathology.

These findings indicate that steeper delay discounting may not be a risk factor for psychiatric disorders. Instead, steeper delay discounting may be a consequence of psychiatric disorders, appearing as or after the disorder develops. It is also possible that associations between delay discounting and risk of psychopathology are not yet apparent at this developmental stage given that the brain’s reward system and prefrontal cortex undergo substantial changes at puberty and in later stages of development (Somerville and Casey 2010). Importantly, given the associations between delay discounting and sociodemographic variables at this age, it is possible that the observed cross-sectional differences in delay discounting behavior in other studies are at least partially due to socioeconomic and demographic differences between cases and controls. In support of this, we have previously found that education and household income explained a substantially greater proportion of the variance in delay discounting behavior than diagnostic group in a study comparing individuals with alcohol use disorder to healthy controls (Gowin et al. 2019). This is consistent with the present analysis, which found that parental education level had the strongest effect on delay discounting behavior at ages 9-11. Future case-control and prospective studies should therefore carefully assess and control for socioeconomic and demographic variables of interest.

Few studies have assessed delay discounting behavior across the lifespan, although there is some evidence that delay discounting varies as a function of age. One study found that delay discounting was steepest in children and adults over age 85 as compared to other age groups (Göllner et al. 2017). Another, study looking at children age 6-19 found that discounting was shallowest in adolescents age 13-17 and steepest in children age 6-12 (Scheres et al. 2014). In the NCANDA study, a study of 831 adolescents age 12-21 recruited across 5 U.S. sites, delay discounting behavior was steepest in early adolescence and became shallower at older ages (Sullivan et al. 2016). These studies suggest that the children recruited in the ABCD study may also have changes in their delay discounting behavior as they age, but another study with repeated measures across development suggests high test-retest reliability of delay discounting, suggesting that these measurements could be reliable trait indices (Klein et al. 2022). There are some prospective data suggesting that steeper delay discounting at older ages could be predictive of problematic behavior. For example, steeper discounting at age 14 was found to predict problematic drug use at age 16 (Büchel et al. 2017). However, another study found no relationship between delay discounting at age 14-15 and either substance use or trait impulsivity in later adolescence (Isen et al. 2014). Studies prospectively linking steeper discounting behavior to the development of other forms of psychopathology such as mood disorders are lacking.

An interesting additional finding of our analyses relates to the data quality criteria for the delay discounting task (Johnson and Bickel 2008). Compared to those who did not meet quality criteria, children whose performance on the task met criteria were more likely to have a parent with a bachelor’s degree or graduate degree (70.7% vs. 54.6%) and to come from families with a household income above $100,000 per year (47.0% vs. 34.7%). Individuals who met these criteria were also more likely to be White (71.9% vs. 59.9%) and less likely to be Black (8.8% vs. 18.6%). This raises important questions as to whether these criteria tend to exclude lower income and historically marginalized groups.

## 4.2 Limitations

Our study had several important limitations. First, we only looked at cross-sectional associations between family history of psychopathology and delay discounting behavior at one time point. It is possible that the observed associations between family history of psychiatric disorders and delay discounting behavior will change as children age and their brains begin to mature. Analysis of future releases of ABCD data will help determine whether this is the case. Second, ABCD’s assessment of family history of psychiatric disorders is limited in that it relies on parental self-report based on single questions. This may lead to false positives, with parents misidentifying relatives as having had a psychiatric disorder. False negatives may also occur due to both the limited scope of the prompts (e.g. an uncle may have had delusions of reference but not paranoid delusions, and these are not assessed in any of the prompts) or because a parent is unaware of certain events (e.g. they may have never been told about a family member’s suicide attempt). A more comprehensive method of assessing family history of psychiatric disorders would have been preferable, but was probably beyond the scope of the ABCD study given the large number of other assessments administered. Nevertheless, the lack of any associations between these items and delay discounting behavior in such a large sample of children provides compelling evidence against family history of psychopathology having strong associations with delay discounting behavior at this age. Third, as mentioned above, delay discounting data for most participants did not meet the data quality criteria for the delay discounting task. Despite this, results were similar in our analyses within the subsample of participants whose data met these quality criteria.

## 4.3 Conclusions

Our results suggest that delay discounting behavior in childhood may not be associated with risk of future psychopathology. Rather, individuals at risk for mental illness may only experience changes in delay discounting in later in adolescence or these changes may occur when psychopathology emerges. In either case, evidence from this study suggests that at this age, socioeconomic and demographic factors have a larger impact on delay discounting behavior than liability for psychopathology. Longitudinal follow up of this cohort will help clarify whether delay discounting behavior predicts future psychopathology and the degree to which delay discounting behavior changes over development.

## Data Availability

All data produced are available from the NIMH Data Archive.

https://nda.nih.gov/

## Acknowledgements

Data used in the preparation of this article were obtained from the Adolescent Brain Cognitive DevelopmentSM (ABCD) Study (https://abcdstudy.org), held in the NIMH Data Archive (NDA). This is a multisite, longitudinal study designed to recruit more than 10,000 children age 9-10 and follow them over 10 years into early adulthood. The ABCD Study® is supported by the National Institutes of Health and additional federal partners under award numbers U01DA041048, U01DA050989, U01DA051016, U01DA041022, U01DA051018, U01DA051037, U01DA050987, U01DA041174, U01DA041106, U01DA041117, U01DA041028, U01DA041134, U01DA050988, U01DA051039, U01DA041156, U01DA041025, U01DA041120, U01DA051038, U01DA041148, U01DA041093, U01DA041089, U24DA041123, U24DA041147. A full list of supporters is available at https://abcdstudy.org/federal-partners.html. A listing of participating sites and a complete listing of the study investigators can be found at https://abcdstudy.org/consortium_members/. ABCD consortium investigators designed and implemented the study and/or provided data but did not necessarily participate in the analysis or writing of this report. This manuscript reflects the views of the authors and may not reflect the opinions or views of the NIH or ABCD consortium investigators.

## Author Contributions

This study was conceived and designed by MES and JLG. MS conducted the data analyses and prepared the figures. The initial draft of the manuscript was written by MES. The study design was shared by all authors and all authors revised the paper for intellectual content and approved the final version to be published.

