## Supplemental Materials for "Delay Discounting and Family History of Psychopathology in Children Ages 9-11: Results from the ABCD Study"

**Figure S1**

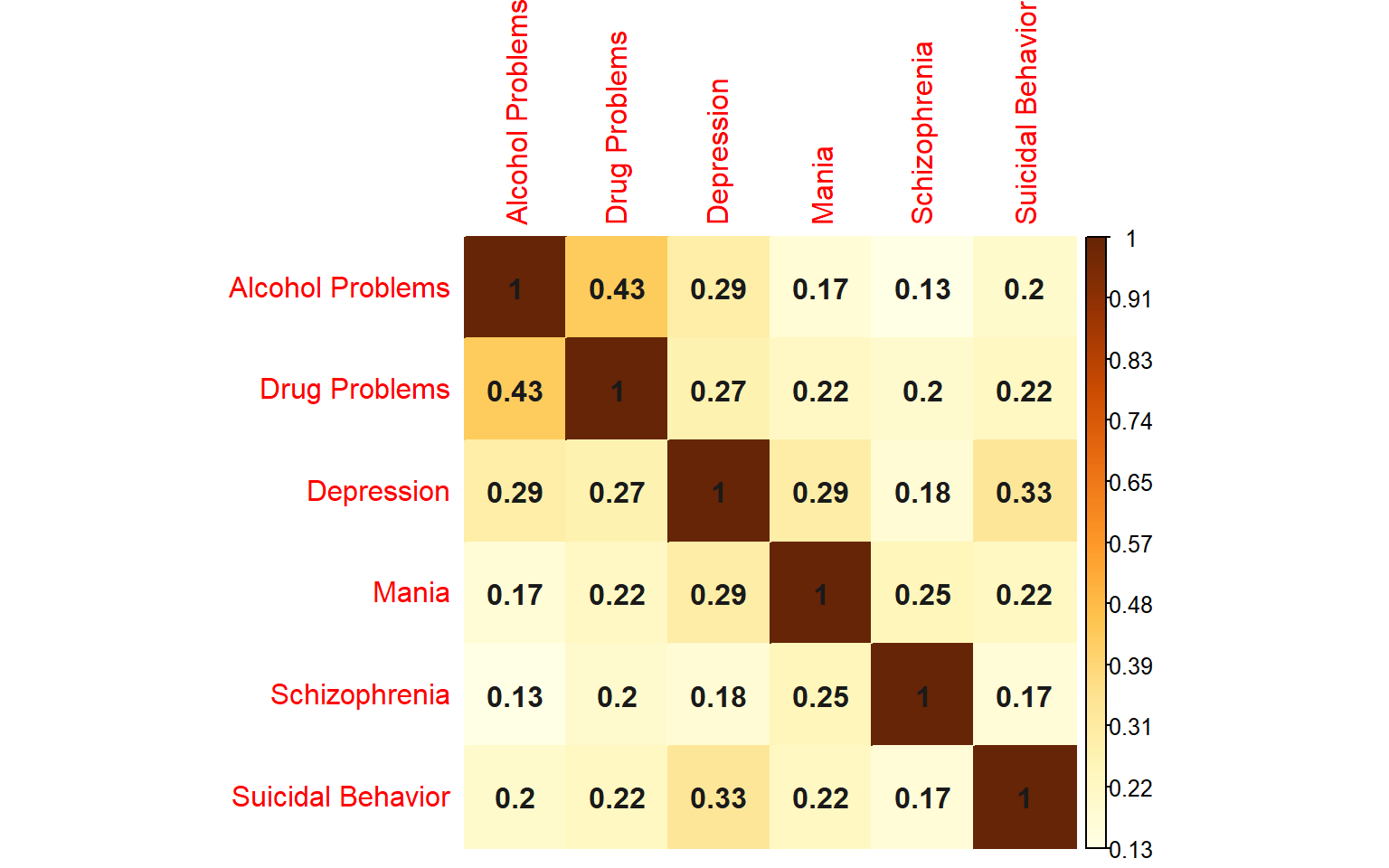

Spearman correlations between family pattern density scores. All correlations were highly significant (p < 0.001). The magnitudes were medium to large.

**Figure S2**

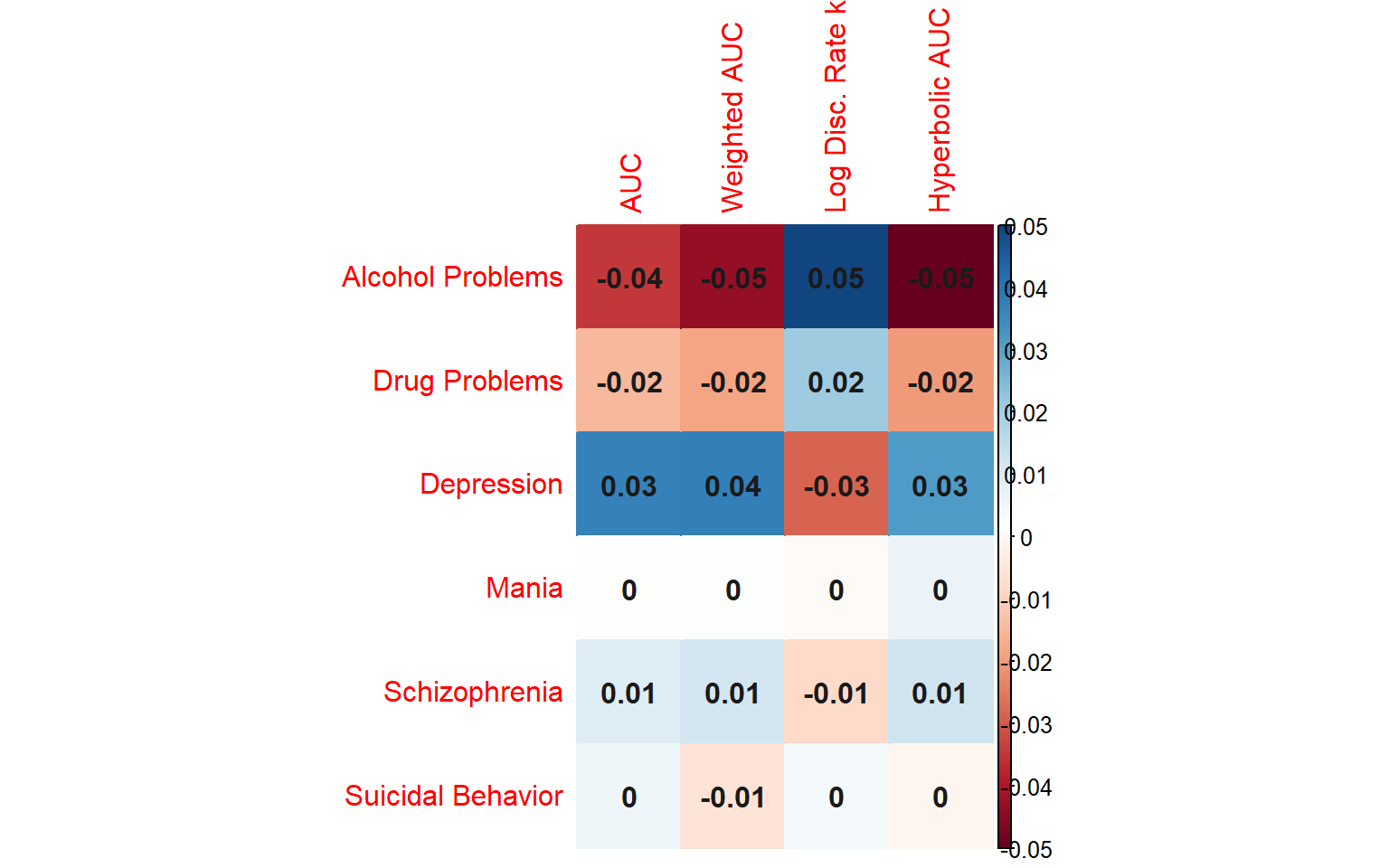

Spearman correlations between family pattern density scores and delay discounting measures in the sample that met quality criteria for the delay discounting task (N=4364). Family history of alcohol problems, drug problems, mania, schizophrenia, and suicide attempt or completion were measured using family pattern density scores. Correlations were small in magnitude, where our primary measure of delay discounting (AUC) had values ranging from ranging from -0.04 to 0.03.

**Figure S3**

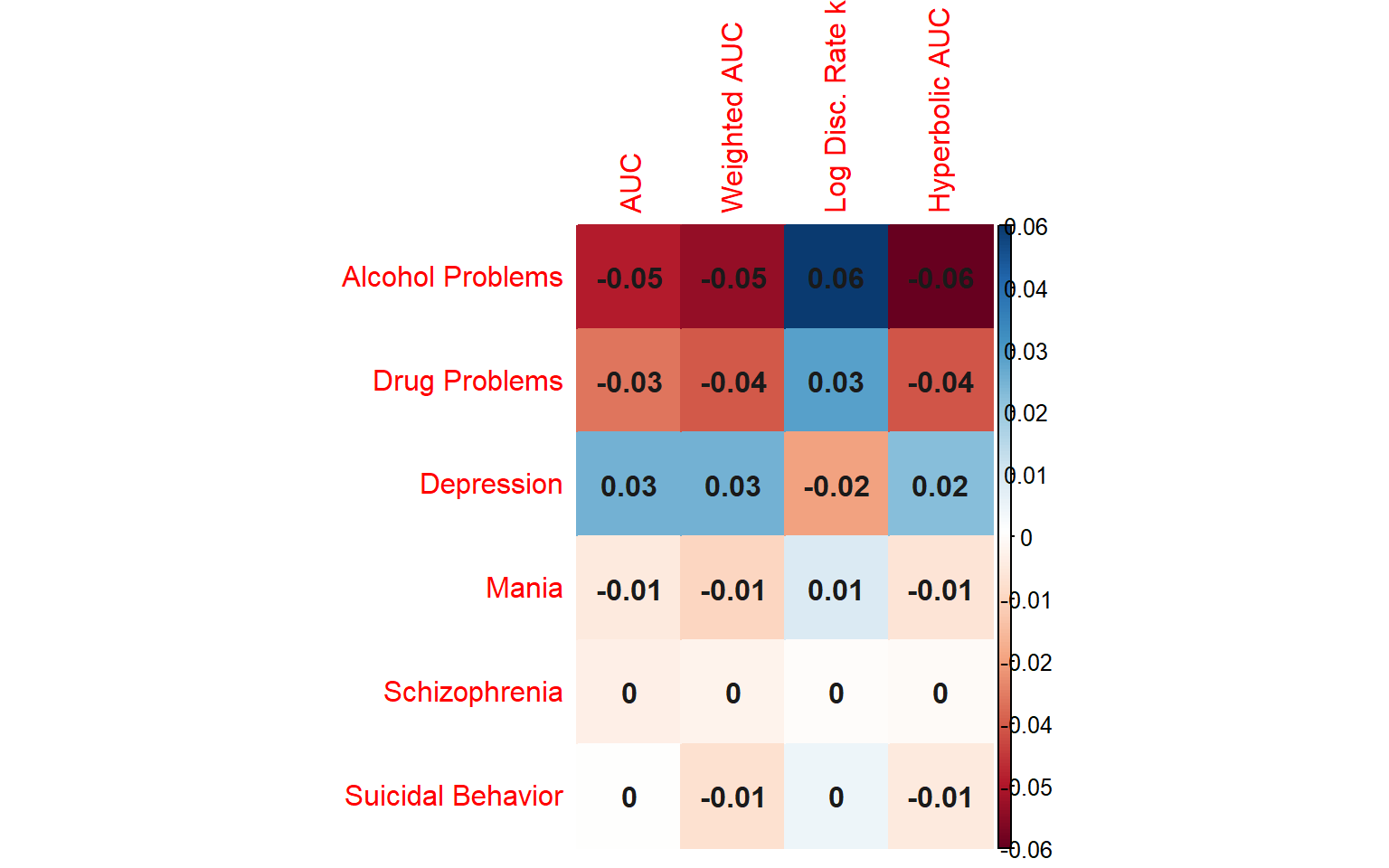

Correlations between family history density scores and delay discounting measures in the sample that met quality criteria for the delay discounting task (N=4364). Family history of alcohol problems, drug problems, mania, schizophrenia, and suicide attempt or completion were measured using family history density scores. Correlations were small in magnitude, where our primary measure of delay discounting (AUC) had values ranging from ranging from -0.05 to 0.03.

**Figure S4**

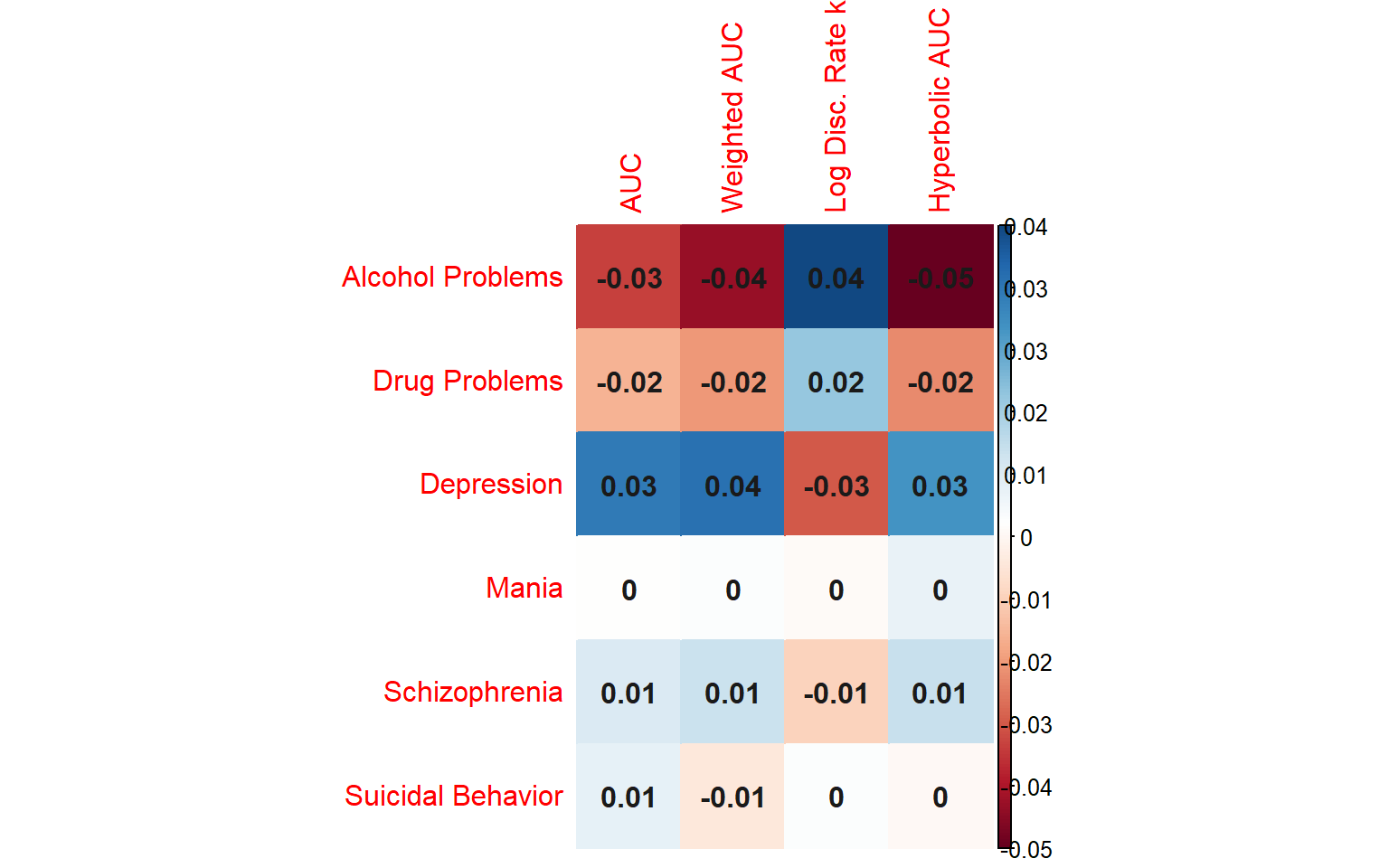

Correlations between family patterns analysis scores and delay discounting measures in the sample that met quality criteria for the delay discounting task (N=4364). Family history of alcohol problems, drug problems, mania, schizophrenia, and suicide attempt or completion were measured using family pattern analysis scores. Correlations were small in magnitude, where our primary measure of delay discounting (AUC) had values ranging from ranging from -0.03 to 0.03.

**Figure S5**

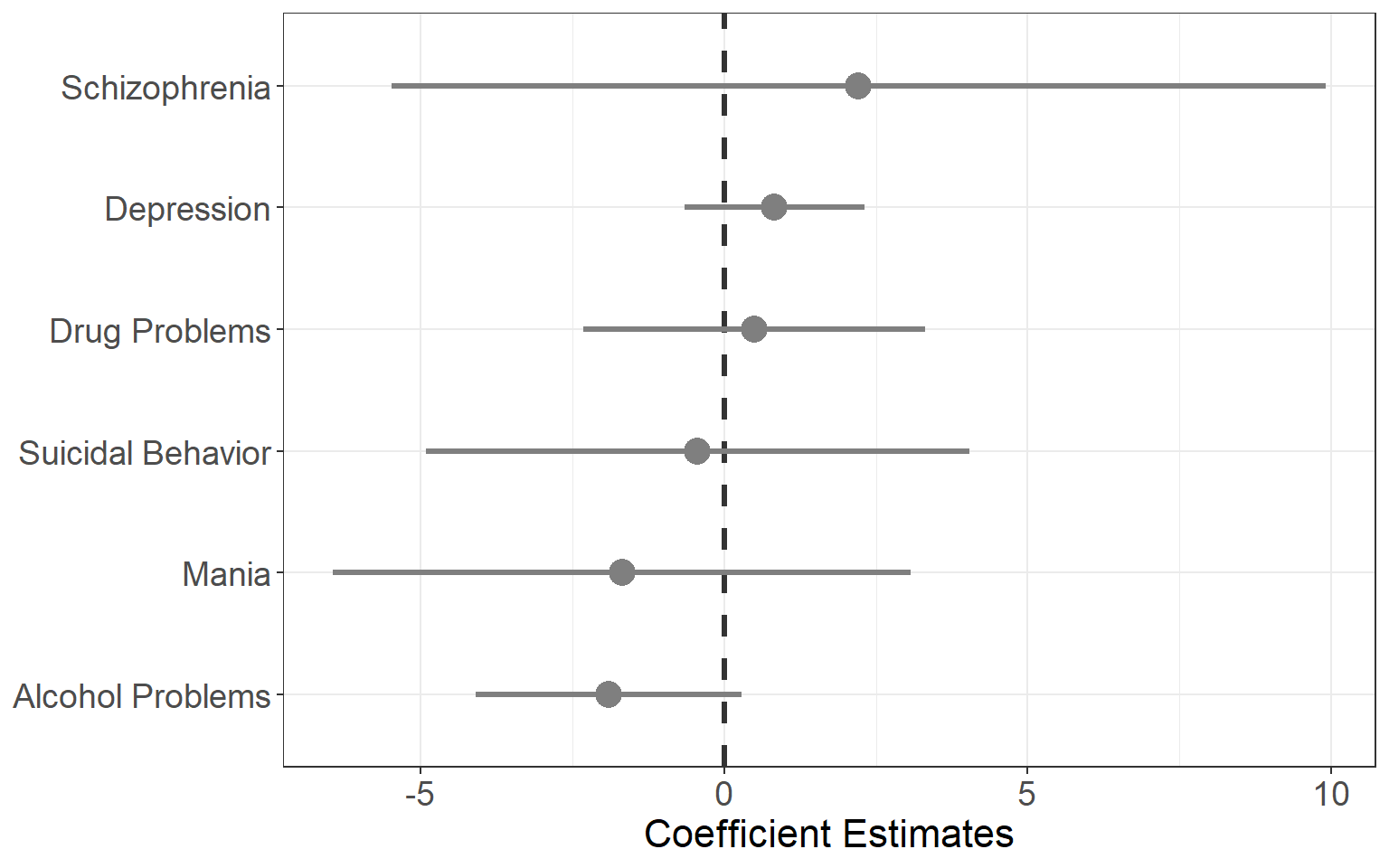

Coefficient estimates from adjusted mixed effects models examining the association between family history density scores and delay discounting behavior (area under the curve) in the sample that met quality criteria for the delay discounting task (N=4364). Mixed effects models did not find any significant associations between family history of psychiatric disorders and delay discounting behavior when adjusted for socioeconomic and demographic variables.

**Figure S6**

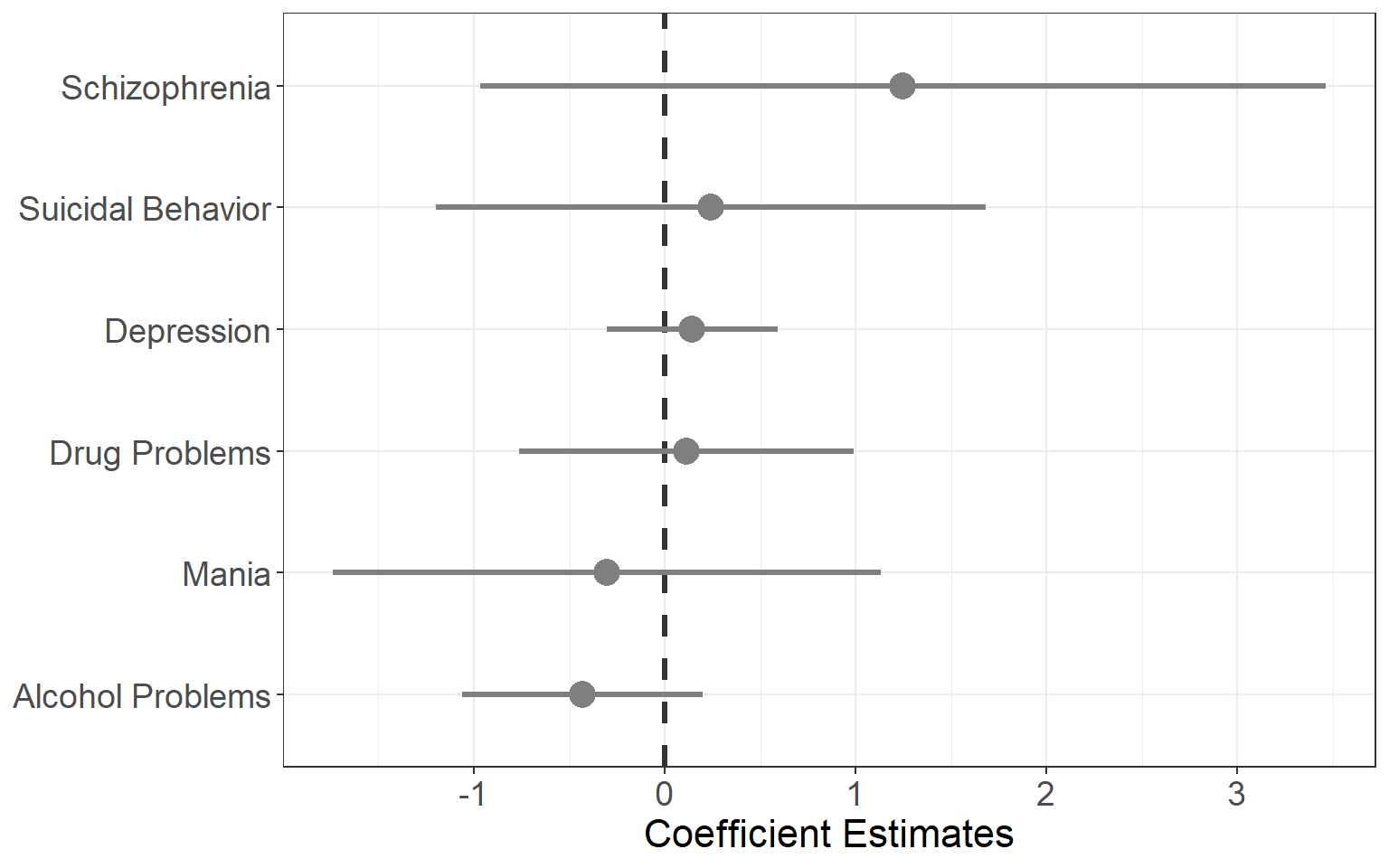

Coefficient estimates from adjusted mixed effects models examining the association between family patterns analysis scores and delay discounting behavior (area under the curve) in the sample that met quality criteria for the delay discounting task (N=4364). Mixed effects models did not find any significant associations between family history of psychiatric disorders and delay discounting behavior when adjusted for socioeconomic and demographic variables.

**Figure S7**

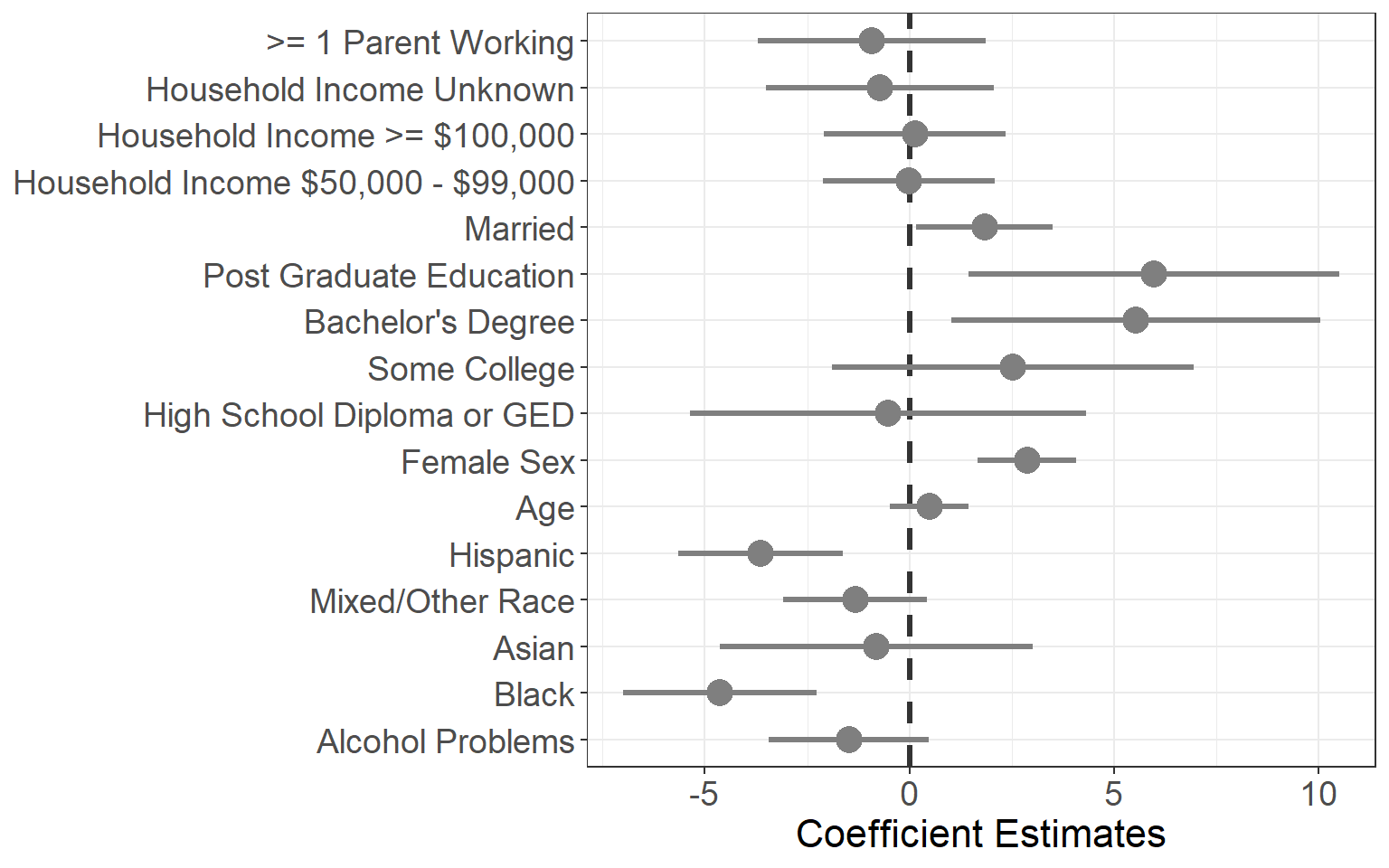

Full mixed effects model examining the association between family pattern density of alcohol problems and delay discounting behavior (area under the curve) in the sample that met quality criteria for the delay discounting task (N=4364). The model did not find a significant association between family history of alcohol problems and delay discounting behavior when adjusted for socioeconomic and demographic variables.

**Figure S8**

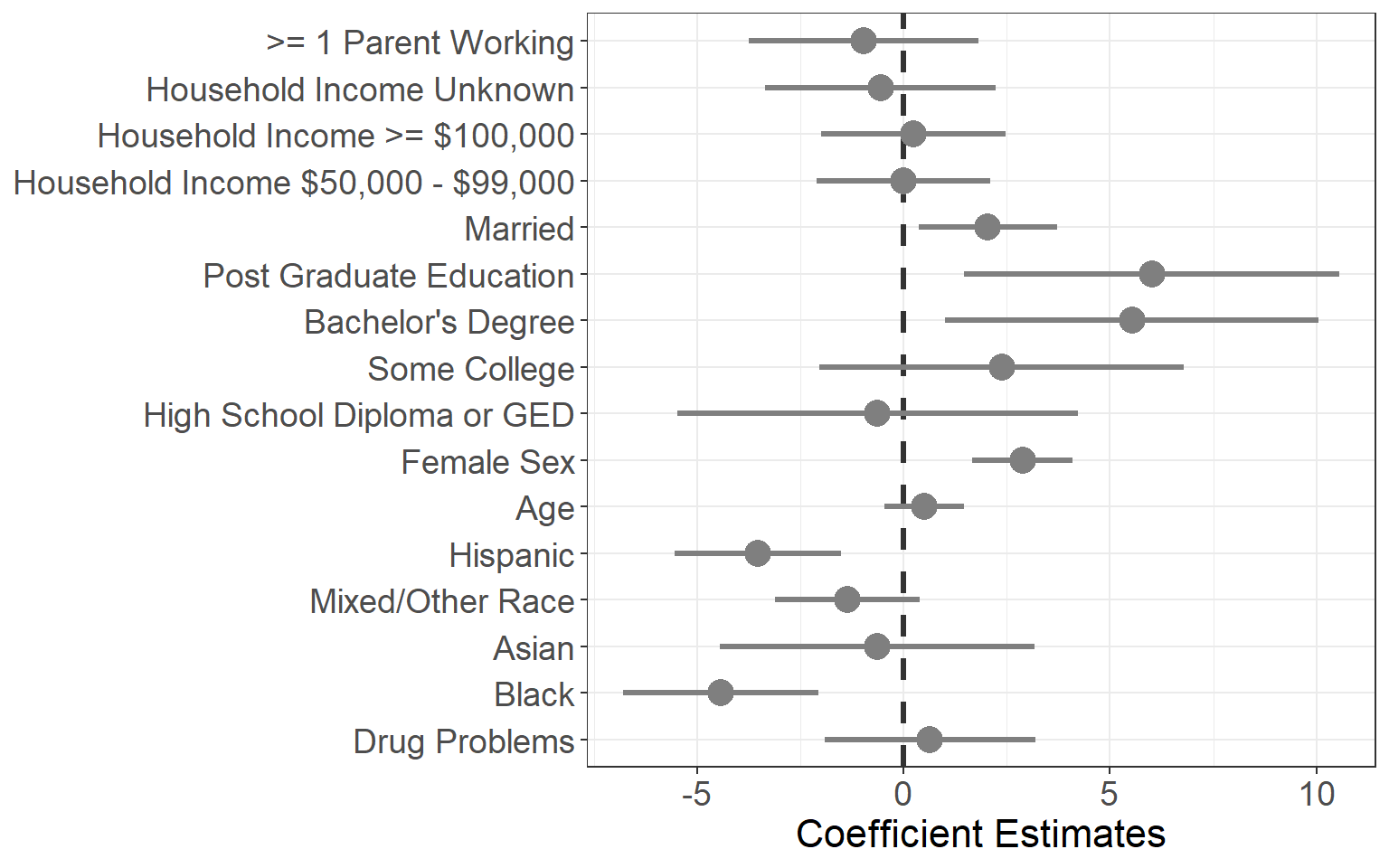

Full mixed effects model examining the association between family pattern density of drug problems and delay discounting behavior (area under the curve) in the sample that met quality criteria for the delay discounting task (N=4364). The model did not find a significant association between family history of drug problems and delay discounting behavior when adjusted for socioeconomic and demographic variables.

**Figure S9**

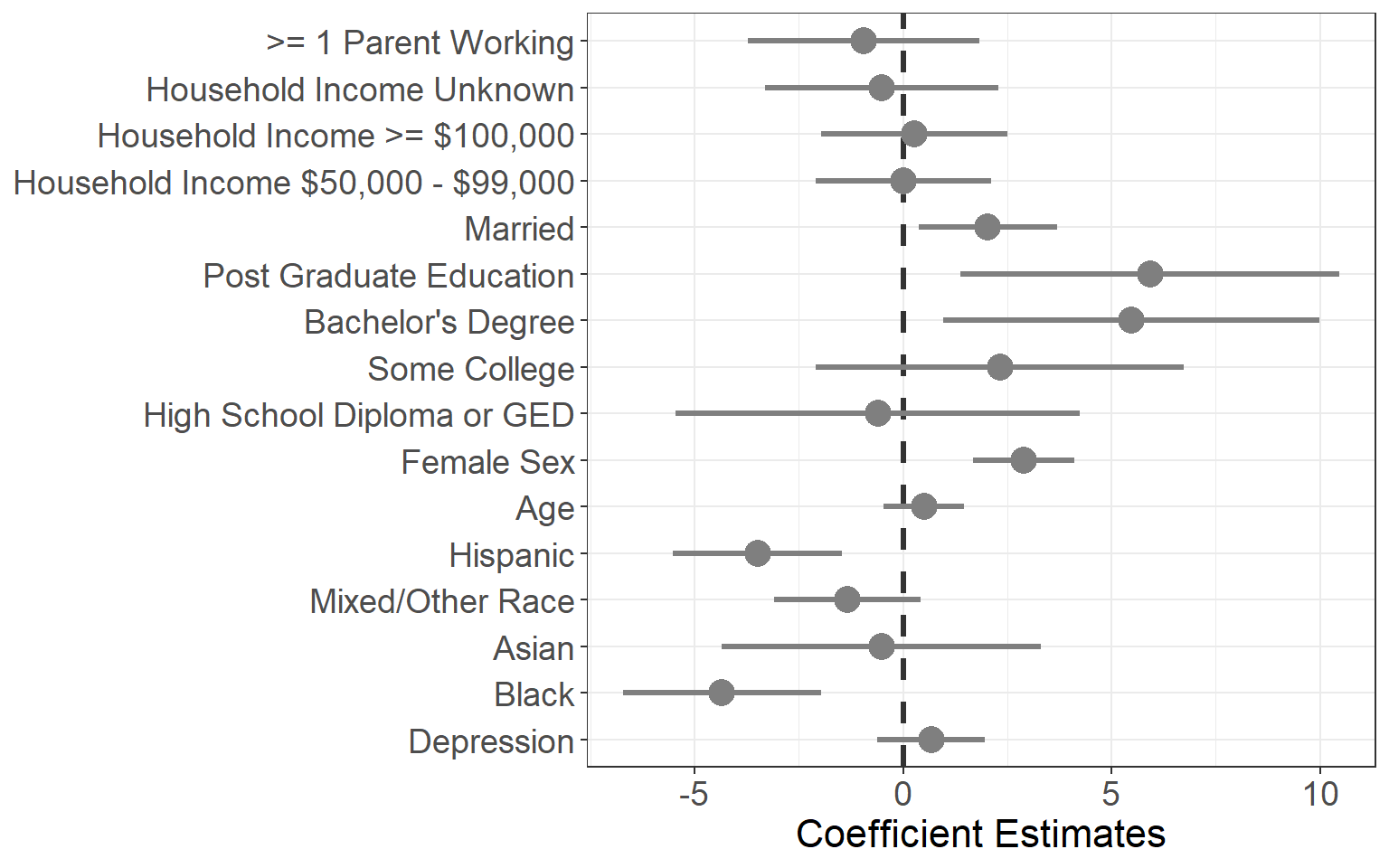

Full mixed effects model examining the association between family pattern density of depression and delay discounting behavior (area under the curve) in the sample that met quality criteria for the delay discounting task (N=4364). The model did not find a significant association between family history of depression and delay discounting behavior when adjusted for socioeconomic and demographic variables.

**Figure S10**

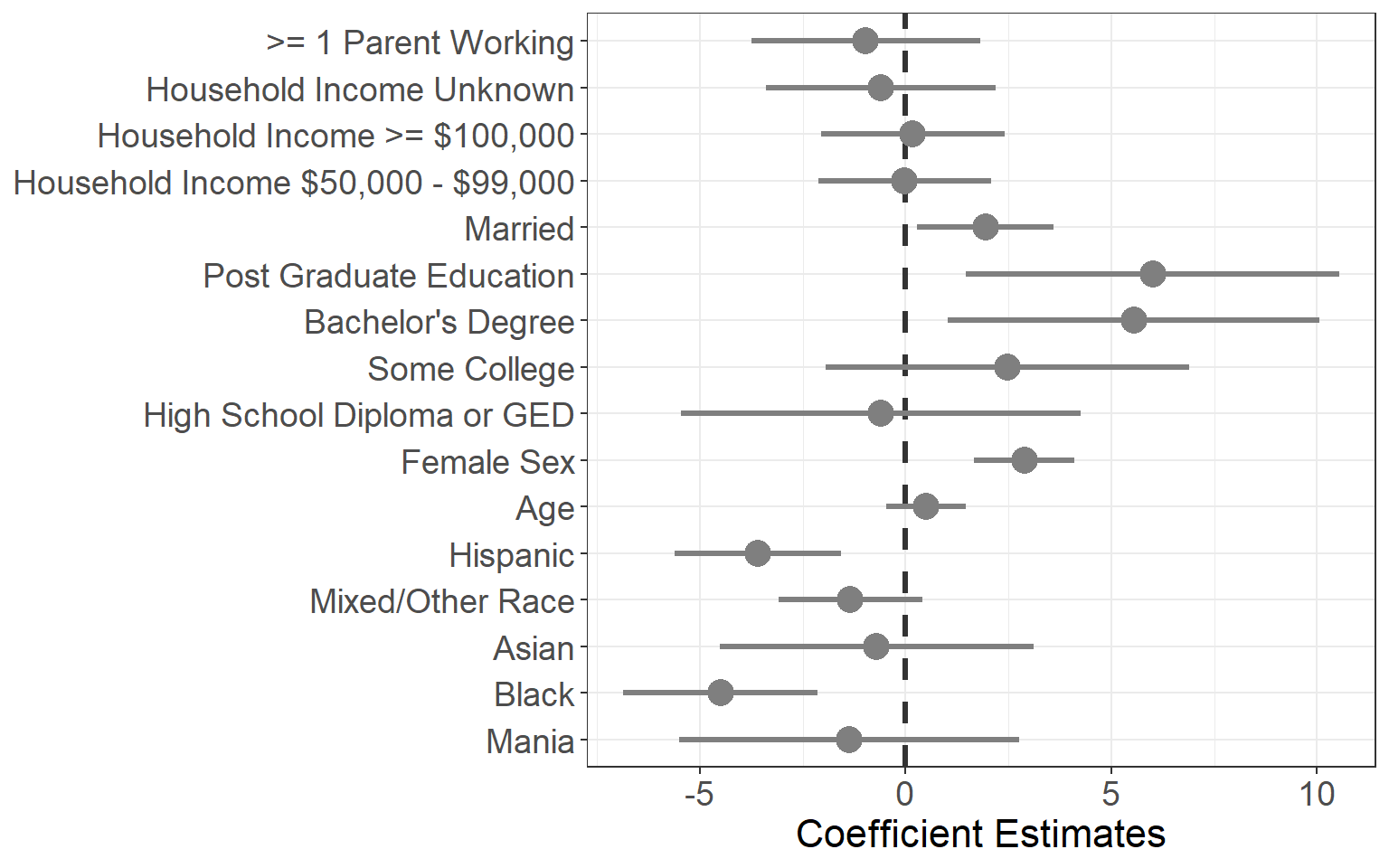

Full mixed effects model examining the association between family pattern density of mania and delay discounting behavior (area under the curve) in the sample that met quality criteria for the delay discounting task (N=4364). The model did not find a significant association between family history of mania and delay discounting behavior when adjusted for socioeconomic and demographic variables.

**Figure S11**

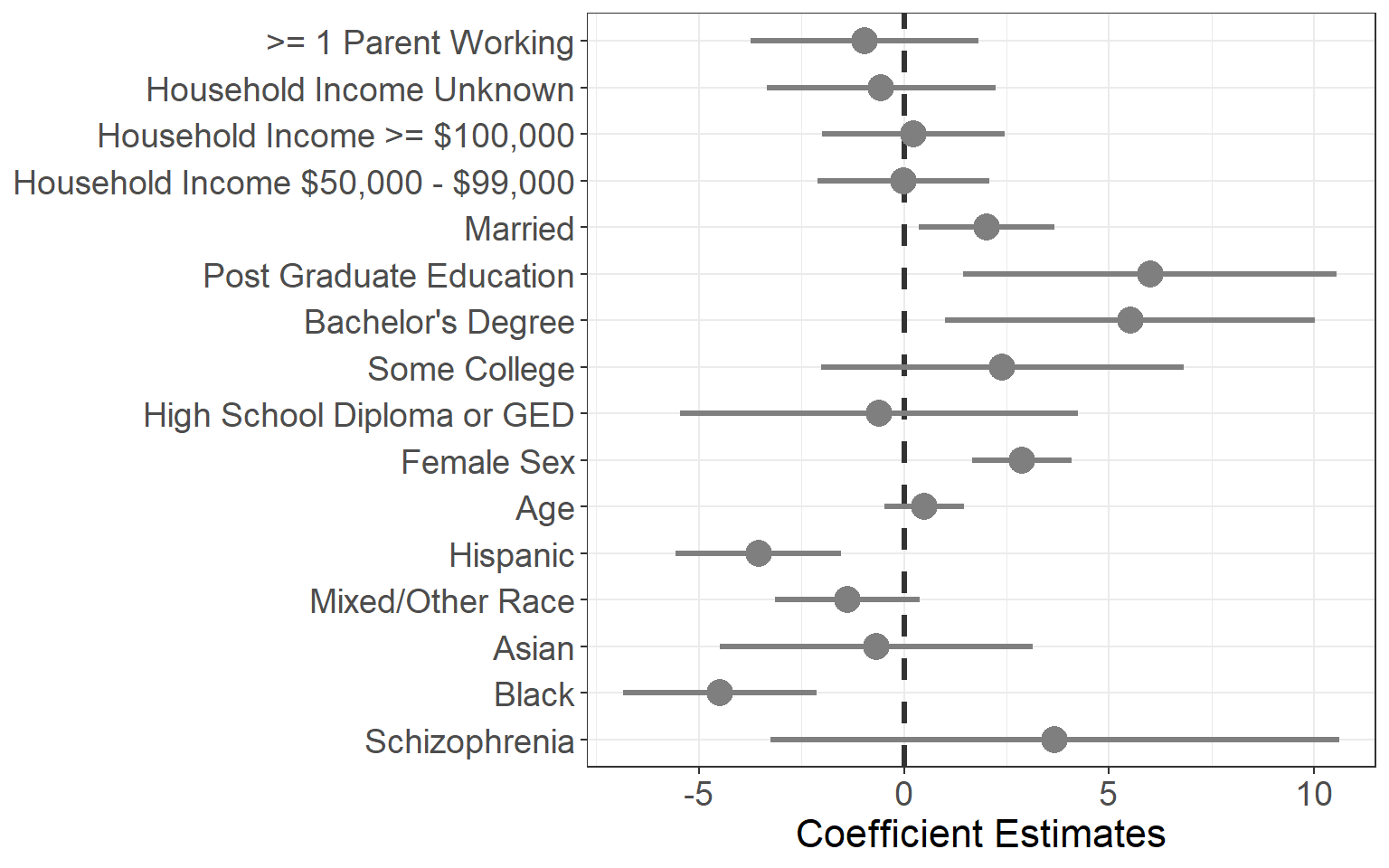

Full mixed effects model examining the association between family pattern density of schizophrenia and delay discounting behavior (area under the curve) in the sample that met quality criteria for the delay discounting task (N=4364). The model did not find a significant association between family history of schizophrenia and delay discounting behavior when adjusted for socioeconomic and demographic variables.

**Figure S12**

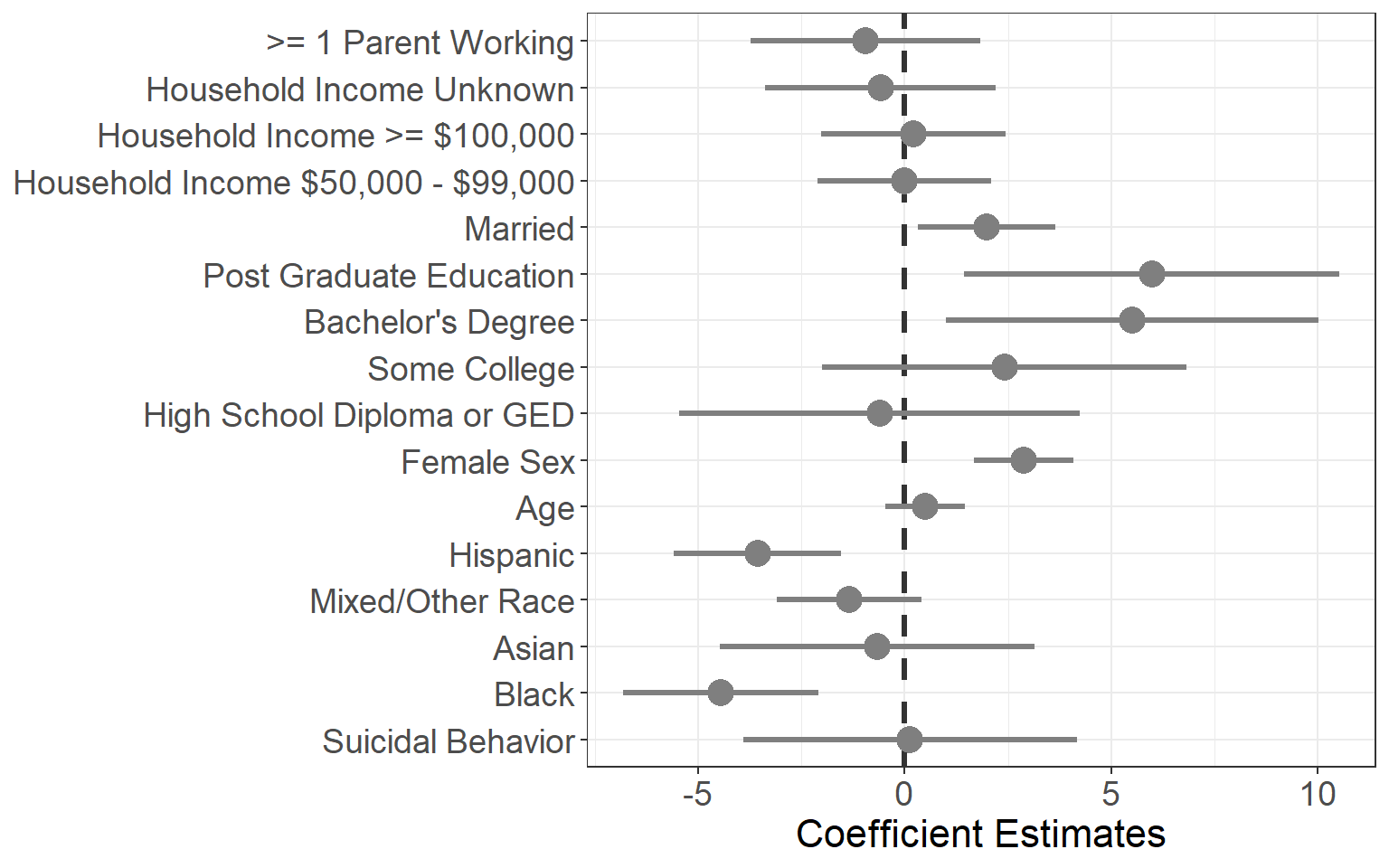

Full mixed effects model examining the association between family pattern density of suicidal behavior and delay discounting behavior (area under the curve) in the sample that met quality criteria for the delay discounting task (N=4364). The model did not find a significant association between family history of suicidal behavior and delay discounting behavior when adjusted for socioeconomic and demographic variables.

**Table S1**

| Predictors | Estimates | 95% CI | p |
| --- | --- | --- | --- |
| Family History of Alcohol Problems | -1.55 | -2.88, -0.22 | 0.022 |
| **Black** | **-7.02** | **-8.48, -5.55** | **<0.001** |
| Asian | -1.69 | -4.59, 1.21 | 0.253 |
| **Mixed/Other Race** | **-2.10** | **-3.33, -0.88** | **0.001** |
| **Hispanic** | **-3.88** | **-5.28, -2.49** | **<0.001** |
| Age | 0.45 | -0.24, 1.13 | 0.199 |
| **Female** | **2.66** | **1.80, 3.52** | **<0.001** |
| High School Diploma or GED | 0.84 | -1.79, 3.46 | 0.532 |
| Some College | 2.13 | -0.27, 4.54 | 0.082 |
| **Bachelor’s Degree** | **5.14** | **2.60, 7.69** | **<0.001** |
| **Post Graduate Education** | **6.29** | **3.71, 8.86** | **<0.001** |
| Married | 1.30 | 0.14, 2.45 | 0.028 |
| Household Income $50,000 - $99,000 | 1.70 | 0.30, 3.10 | 0.018 |
| Household Income ≥ $100,000 | 1.20 | -0.34, 2.73 | 0.126 |
| Household Income Unknown | 0.65 | -1.16, 2.46 | 0.482 |
| ≥1 Parent Working | 0.83 | -0.83, 2.49 | 0.326 |
| Random Effects |  |  |  |
| σ | 453.57 |  |  |
| τ_00_ Family | 55.19 |  |  |
| τ_00_ Site | 0.74 |  |  |
| ICC | 0.11 |  |  |
| N_Family_ | 8944 |  |  |
| N_Site_ | 22 |  |  |
| Observations | 10788 |  |  |
| Marginal R^2^ | 0.049 |  |  |
| Conditional R^2^ | 0.152 |  |  |

Full mixed effects model examining the association between family pattern density of alcohol problems and delay discounting behavior (area under the curve) in the total sample. The model did not find a significant association between family history of alcohol problems and delay discounting behavior when adjusted for socioeconomic and demographic variables and applying Bonferroni correction for six comparisons.

**Table S2**

| Predictors | Estimates | 95% CI | p |
| --- | --- | --- | --- |
| Family History of Drug Problems | -1.43 | -3.04, 0.19 | 0.084 |
| **Black** | **-6.90** | **-8.37, -5.44** | **<0.001** |
| Asian | -1.60 | -4.49, 1.30 | 0.280 |
| **Mixed/Other Race** | **-2.09** | **-3.32, -0.86** | **0.001** |
| **Hispanic** | **-3.87** | **-5.27, -2.48** | **<0.001** |
| Age | 0.45 | -0.23, 1.13 | 0.197 |
| **Female** | **2.66** | **1.80, 3.52** | **<0.001** |
| High School Diploma or GED | 0.86 | -1.76, 3.48 | 0.521 |
| Some College | 2.13 | -0.28, 4.54 | 0.083 |
| **Bachelor’s Degree** | **5.11** | **2.56, 7.65** | **<0.001** |
| **Post Graduate Education** | **6.28** | **3.70, 8.85** | **<0.001** |
| Married | 1.32 | 0.16, 2.48 | 0.025 |
| Household Income $50,000 - $99,000 | 1.68 | 0.28, 3.09 | 0.019 |
| Household Income ≥ $100,000 | 1.17 | -0.36, 2.71 | 0.134 |
| Household Income Unknown | 0.67 | -1.14, 2.48 | 0.469 |
| ≥ 1 Parent Working | 0.81 | -0.85, 2.47 | 0.340 |
| Random Effects |  |  |  |
| σ | 453.93 |  |  |
| τ_00_ Family | 53.9 |  |  |
| τ_00_ Site | 0.79 |  |  |
| ICC | 0.11 |  |  |
| N_Family_ | 8944 |  |  |
| N_Site_ | 22 |  |  |
| Observations | 10788 |  |  |
| Marginal R^2^ | 0.049 |  |  |
| Conditional R^2^ | 0.151 |  |  |

Full mixed effects model examining the association between family pattern density of drug problems and delay discounting behavior (area under the curve) in the full sample. The model did not find a significant association between family history of drug problems and delay discounting behavior when adjusted for socioeconomic and demographic variables.

**Table S3**

| Predictors | Estimates | 95% CI | p |
| --- | --- | --- | --- |
| Family History of Depression | 0.13 | -0.80, 1.06 | 0.787 |
| **Black** | **-6.78** | **-8.26, -5.31** | **<0.001** |
| Asian | -1.47 | -4.37, 1.43 | 0.322 |
| **Mixed/Other Race** | **-2.11** | **-3.34, -0.88** | **0.001** |
| **Hispanic** | **-3.77** | **-5.16, -2.37** | **<0.001** |
| Age | 0.44 | -0.24, 1.13 | 0.203 |
| **Female Sex** | **2.66** | **1.80, 3.52** | **<0.001** |
| High School Diploma or GED | 0.77 | -1.85, 3.40 | 0.563 |
| Some College | 2.00 | -0.41, 4.40 | 0.104 |
| **Bachelor’s Degree** | **5.09** | **2.55, 7.64** | **<0.001** |
| **Post Graduate Education** | **6.29** | **3.71, 8.87** | **<0.001** |
| Married | 1.47 | 0.32, 2.62 | 0.012 |
| Household Income $50,000 - $99,000 | 1.72 | 0.31, 3.12 | 0.017 |
| Household Income ≥ $100,000 | 1.26 | -0.27, 2.79 | 0.108 |
| Household Income Unknown | 0.76 | -1.06, 2.57 | 0.412 |
| ≥Parent Working | 0.86 | -0.80, 2.52 | 0.309 |
| Random Effects |  |  |  |
| σ | 454.03 |  |  |
| τ_00_ Family | 53.94 |  |  |
| τ_00_ Site | 0.78 |  |  |
| ICC | 0.11 |  |  |
| N_Family_ | 8944 |  |  |
| N_Site_ | 22 |  |  |
| Observations | 10788 |  |  |
| Marginal R^2^ | 0.049 |  |  |
| Conditional R^2^ | 0.151 |  |  |

Full mixed effects model examining the association between family pattern density of depression and delay discounting behavior (area under the curve) in the full sample. The model did not find a significant association between family history of depression and delay discounting behavior when adjusted for socioeconomic and demographic variables.

**Table S4**

| Predictors | Estimates | 95% CI | p |
| --- | --- | --- | --- |
| Family History of Mania | -2.91 | -5.69, -0.14 | 0.040 |
| **Black** | **-6.88** | **-8.35, -5.42** | **<0.001** |
| Asian | -1.58 | -4.47, 1.32 | 0.285 |
| **Mixed/Other Race** | **-2.09** | **-3.31, -0.86** | **0.001** |
| **Hispanic** | **-3.86** | **-5.26, -2.47** | **<0.001** |
| Age | 0.45 | -0.24, 1.13 | 0.201 |
| **Female** | **2.64** | **1.79, 3.50** | **<0.001** |
| High School Diploma or GED | 0.81 | -1.81, 3.43 | 0.544 |
| Some College | 2.12 | -0.29, 4.52 | 0.085 |
| **Bachelor’s Degree** | **5.17** | **2.62, 7.71** | **<0.001** |
| **Post Graduate Education** | **6.35** | **3.78, 8.92** | **<0.001** |
| Married | 1.39 | 0.24, 2.54 | 0.018 |
| Household Income $50,000 - $99,000 | 1.66 | 0.26, 3.07 | 0.020 |
| Household Income ≥ $100,000 | 1.19 | -0.34, 2.73 | 0.127 |
| Household Income Unknown | 0.69 | -1.12, 2.51 | 0.452 |
| ≥ 1 Parent Working | 0.77 | -0.89, 2.43 | 0.365 |
| Random Effects |  |  |  |
| σ | 454.09 |  |  |
| τ_00_ Family | 53.65 |  |  |
| τ_00_ Site | 0.82 |  |  |
| ICC | 0.11 |  |  |
| N_Family_ | 8944 |  |  |
| N_Site_ | 22 |  |  |
| Observations | 10788 |  |  |
| Marginal R^2^ | 0.049 |  |  |
| Conditional R^2^ | 0.151 |  |  |

Full mixed effects model examining the association between family pattern density of mania and delay discounting behavior (area under the curve) in the full sample. The model did not find a significant association between family history of mania and delay discounting behavior when adjusted for socioeconomic and demographic variables.

**Table S5**

| Predictors | Estimates | 95% CI | p |
| --- | --- | --- | --- |
| Family History of Schizophrenia | 0.62 | -3.74, 4.98 | 0.780 |
| **Black** | **-6.81** | **-8.27, -5.35** | **<0.001** |
| Asian | -1.50 | -4.39, 1.40 | 0.310 |
| **Mixed/Other Race** | **-2.12** | **-3.34, -0.89** | **0.001** |
| **Hispanic** | **-3.78** | **-5.17, -2.39** | **<0.001** |
| Age | 0.45 | -0.24, 1.13 | 0.202 |
| **Female** | **2.66** | **1.80, 3.51** | **<0.001** |
| High School Diploma or GED | 0.78 | -1.84, 3.40 | 0.560 |
| Some College | 2.01 | -0.40, 4.41 | 0.102 |
| **Bachelor’s Degree** | **5.11** | **2.56, 7.65** | **<0.001** |
| **Post Graduate Education** | **6.30** | **3.73, 8.88** | **<0.001** |
| Married | 1.46 | 0.31, 2.61 | 0.013 |
| Household Income $50,000 - $99,000 | 1.72 | 0.31, 3.12 | 0.017 |
| Household Income ≥ $100,000 | 1.26 | -0.28, 2.79 | 0.108 |
| Household Income Unknown | 0.76 | -1.06, 2.57 | 0.414 |
| ≥1 Parent Working | -0.86 | -0.80, 2.52 | 0.308 |
| Random Effects |  |  |  |
| σ | 454.03 |  |  |
| τ_00_ Family | 53.93 |  |  |
| τ_00_ Site | 0.79 |  |  |
| ICC | 0.11 |  |  |
| N_Family_ | 8944 |  |  |
| N_Site_ | 22 |  |  |
| Observations | 10788 |  |  |
| Marginal R^2^ | 0.049 |  |  |
| Conditional R^2^ | 0.151 |  |  |

Full mixed effects model examining the association between family pattern density of schizophrenia and delay discounting behavior (area under the curve) in the sample meeting the Johnson and Bickel criteria. The model did not find a significant association between family history of schizophrenia and delay discounting behavior when adjusted for socioeconomic and demographic variables.

**Table S6**

| Predictors | Estimates | 95% CI | p |
| --- | --- | --- | --- |
| Family History of Suicidal Behavior | -0.04 | -2.82, 2.73 | 0.975 |
| **Black** | **-6.81** | **-8.28, -5.35** | **<0.001** |
| Asian | -1.50 | -4.39, 1.39 | 0.310 |
| **Mixed/Other Race** | **-2.11** | **-3.34, -0.89** | **0.001** |
| **Hispanic** | **-3.79** | **-5.18, -2.39** | **<0.001** |
| Age | 0.44 | -0.24, 1.13 | 0.204 |
| **Female Sex** | **2.66** | **1.80, 3.52** | **<0.001** |
| High School Diploma or GED | 0.78 | -1.84, 3.40 | 0.558 |
| Some College | 2.02 | -0.39, 4.42 | 0.100 |
| **Bachelor’s Degree** | **5.11** | **2.57, 7.65** | **<0.001** |
| **Post Graduate Education** | **6.30** | **3.73, 8.88** | **<0.001** |
| Married | 1.45 | 0.31, 2.60 | 0.013 |
| Household Income $50,000 - $99,000 | 1.71 | 0.31, 3.12 | 0.017 |
| Household Income ≥ $100,000 | 1.25 | -0.29, 2.78 | 0.111 |
| Household Income Unknown | 0.75 | -1.07, 2.56 | 0.420 |
| ≥1 Parent Working | 0.85 | -0.81, 2.51 | 0.316 |
| Random Effects |  |  |  |
| σ | 454.00 |  |  |
| τ_00_ Family | 53.97 |  |  |
| τ_00_ Site | 0.79 |  |  |
| ICC | 0.11 |  |  |
| N_Family_ | 8944 |  |  |
| N_Site_ | 22 |  |  |
| Observations | 10788 |  |  |
| Marginal R^2^ | 0.049 |  |  |
| Conditional R^2^ | 0.151 |  |  |

Full mixed effects model examining the association between family pattern density of suicidal behavior and delay discounting behavior (area under the curve) in the full sample. The model did not find a significant association between family history of suicidal behavior and delay discounting behavior when adjusted for socioeconomic and demographic variables.
